# Prevalence of asymptomatic glioma and implications for survival

**DOI:** 10.1101/2020.04.27.20080564

**Authors:** Paula Province Warren, Mina Lobbous, Noah C. Peeri, Zachary J. Thompson, Reid C. Thompson, Jeffrey J. Olson, Renato V. LaRocca, Sajeel A. Chowdhary, Mark D. Anderson, Michael A. Vogelbaum, James M. Markert, Louis B. Nabors, Kathleen M. Egan

## Abstract

**Background:** Brain tumors can present as focal neurologic deficits (reflecting the tumor location) or generalized symptoms due to increased intracranial pressure. Occasionally, brain tumors can be found incidentally in asymptomatic patients or in patients with unrelated symptoms who undergo brain imaging. The term incidentaloma is used to refer to these imaging abnormalities.

**Objective:** The object of this study was to examine the prevalence and correlates of asymptomatic glioma in a large epidemiological study of brain tumors.

**Methods:** The analysis was based on a large series of patients with glioma (N = 1989) enrolled in a multicenter clinic-based epidemiologic study between 2005 and 2017. Patients were considered asymptomatic from the tumor, and thus as having an incidentally detected glioma (IDG), if the tumor was diagnosed during workup of injury or unrelated medical condition.

**Results:** A total of 32 of 1989 (1.6%) patients were asymptomatic at diagnosis. The leading indication for brain imaging in IDG was non-workplace injuries followed by medical workup for unrelated conditions. IDG was more prevalent in patients younger than 50 years of age (2.6% vs 1.0%). IDG was also more common in patients with low grade gliomas (4.7% for WHO grade II and 1.5% for WHO grade III) vs glioblastomas (0.6% in WHO grade IV).

**Conclusion:** The present data suggest that gliomas may be found incidentally, especially among low grade gliomas. Studies of IDG may be useful as a proxy for early detection of tumor as a means to improve patient survival.

## Introduction

The majority of patients with brain tumors present with either focal or generalized symptoms. Common presenting symptoms include seizures, symptoms of increased intracranial pressure, like headache and/or double vision, or focal neurologic deficits that depend on the intracranial location of the tumor ^1^. Some patients, however, are asymptomatic at the time of presentation and have tumors that are incidentally discovered ^2^. An “incidentaloma” is defined as an imaging abnormality that is detected by chance in patients who undergo imaging for an unrelated medical issue ^3,4^. There has been a rapid rise in incidentaloma detection due to the increase in the number of imaging scans performed as well as improvement in imaging resolution. Common reasons for patients to undergo screening brain scans include trauma or medical workup for an unrelated condition. A 2009 systematic review of 16 studies in 19,559 patients reported the prevalence of incidental neoplastic brain findings in MRI studies to be 0.7%, with the prevalence of glioma at 0.05% ^5^. Diffuse low-grade glioma may be radiologically detectable but clinically silent for more than a decade ^6^.

Advances in molecular profiling and in the understanding of gliomagenesis has allowed for improved diagnostic accuracy and better prognostic markers ^7,8^. Although current recommendations support early surgical resection for low-grade gliomas ^9-13^, management of asymptomatic incidentally diagnosed gliomas remains a matter of debate. There are multiple studies supporting moving away from the “watchful waiting” approach to earlier therapeutic strategies ^14-18^. Pallud *et al* reported that incidental grade II gliomas were progressive in all cases and transformed to symptomatic grade II gliomas at a median interval of 48 months after radiological discovery and had malignant transformation at a median interval of 5.7 years after radiological diagnosis ^4,19^. In addition, the identification of an IDG offers insight into the natural history of the disease prior to the time of neurological presentation.

## Subjects and Methods

### Study Population

The study population was comprised of 1,989 patients with a recent diagnosis of primary glioma (ICD9/10: 191, C71), including glioblastoma (9440-9441) and WHO (World Health Organization) grade II and III gliomas (9382, 9400-01, 9410-11, 9420, 9424-25, 9450-9451)^20,21^, enrolled in an epidemiologic study of glioma ^22^ (“GliomaSE”) between 2005 and 2017. Patients were identified at neuro-oncology and neuro-surgery clinics at major medical centers in the Southeastern United States including Moffitt Cancer Center in Tampa, Florida; Kentuckiana Cancer Center (now Norton Cancer Institute) in Louisville, Kentucky; University of Mississippi Medical Center in Jackson, Mississippi; Emory University in Atlanta, Georgia; University of Alabama at Birmingham in Birmingham, Alabama; Morton Plant Hospital in Tampa, Florida; Vanderbilt University Medical Center in Nashville, Tennessee; Florida Hospital in Orlando, Florida; and Boca Raton Regional Hospital in Boca Raton, Florida). The study was approved by Investigational Review Committees at each participating center and all subjects provided written informed consent. Clinical data including presenting signs and symptoms were collected from neuro-oncology reports. Cause of death and follow-up for vital status were obtained through review of medical records, hospital registries, and a search of the National Death Index.

### Symptoms at Presentation

Medical records for all patients were reviewed for first signs and symptoms of tumor presentation. Symptoms included headache, change in vision or hearing, focal weakness or numbness, memory loss, personality changes, speech difficulties, dizziness or loss of balance, gait or walking difficulties, confusion, and seizure. Patients were considered asymptomatic if the tumor was diagnosed incidentally during work-up for an injury or other non-related medical or surgical conditions. Neuro-oncology notes for each suspected IDG were reviewed by a board-certified neuro-oncologist (PPW). Cases in which events surrounding diagnostic neuroimaging were potentially related to early manifestations of the tumor (for example, a fall with head injury preceded by apparent seizure) were excluded from the IDG category.

### Statistical Analysis

Associations between symptom status at presentation and demographic and clinical factors were assessed with Monte-Carlo estimates of exact *chi-square* test p-values. Logistic regression was used to estimate Odds Ratios (OR) and 95% Confidence Intervals (CI) relating clinical factors to IDG. Multivariable models included terms for tumor grade (Glioblastoma WHO grade IV, WHO grade III, and WHO grade II), gender, and patient age (<50 and >=50 years of age). To evaluate relationships with survival, we selected 1 to 3 symptomatic patients matched to each IDG patient on age at diagnosis (within 5 years), date of diagnosis (within 18 months), and tumor subtype (WHO grade II, III, and IV). Conditional Cox proportional hazards regression was used to estimate the Hazards Ratio (HR) and 95% CIs for IDG with adjustment via matching for age at diagnosis, date of diagnosis, and tumor subtype with observation beginning at first presentation of the tumor.

## Results

A total of 32 of 1989 (1.6%) patients were asymptomatic from the tumor at diagnosis. Circumstances surrounding incidental glioma diagnosis are summarized in **Table 1**. The leading indications were non workplace-related injuries (n=7; examples: patient hit by a falling tree branch), followed by diagnoses made during medical work-up for unrelated diseases or conditions (n=6; examples: sleep apnea; progressive Parkinson’s disease with dementia) and motor vehicle accidents in which the patient was not the driver or judged to be at fault (n=5; example: rear-ended by speeding car). Other categories included pre-operative workup for unrelated CNS surgery (n=1; pre-op for lumbar surgery); surveillance imaging after surgical repair of cerebral aneurysm (n=1, 6 month surveillance scan status-post left middle cerebral artery aneurysm clipping); examination for substance abuse (n=2; example: alcohol withdrawal and fentanyl overdose); infectious disease workup (n=2; viral meningitis and Epstein Barr mononucleosis); hypertension or diabetes workup (n=2; unexplained severe hypertension and uncontrolled diabetes); injury at work (n=2; example: hit in the head by falling box); peripheral neuropathy (n=1; neuropathy due to entrapment of ulnar nerve); unexplained dizziness (n=1; dizziness following brachytherapy for prostate cancer); infertility workup (n=1; related to increased levels of prolactin); and routine eye exam (n=1; papilledema observed during yearly eye exam in a patient with no noted symptoms).

**Table 1.** Circumstances of incidental glioma diagnosis

| Reason for incidental diagnosis | N (%) |
| --- | --- |
| Nonworkplace injury | 7 (21) |
| Follow up for unrelated medical condition | 6 (19) |
| Motor vehicle accident | 5 (16) |
| Preoperative examination | 2 (6) |
| Substance abuse examination | 2 (6) |
| Infectious disease workup | 2 (6) |
| Hypertension and/or diabetes workup | 2 (6) |
| Workplace injury | 2 (6) |
| Peripheral neuropathy workup | 1 (3) |
| Unexplained dizziness | 1 (3) |
| Infertility workup | 1 (3) |
| Routine eye exam | 1 (3) |

Associations between symptom status at presentation and demographic and clinical factors are shown in **Table 2**. Prevalence of IDG was significantly higher in patients under 50 years of age (2.6%) than patients 50 years or older (1.0%). IDG was more common with progressively lower tumor grade: 0.6% for glioblastoma; 1.5% for WHO grade III; and 4.7% for WHO grade II. No significant differences were observed by gender, race, state of residence, education, or hemispheric location of the tumor.

**Table 2.** Descriptive statistics and symptoms at presentation in 1989 glioma patients

| Variable | Level | Symptomatic at Presentation* |  |  |
| --- | --- | --- | --- | --- |
|  |  | Yes<br>(n=1957) | No<br>(n=32) | p-value |
| Age at Diagnosis | <50 | 762 (97.4%) | 20 (2.6%) | **0.01** |
|  | ≥ 50 | 1195 (99.0%) | 12 (1.0%) |  |
| Gender | Female | 826 (98.7%) | 11 (1.3%) | 0.47 |
|  | Male | 1131 (98.2%) | 21 (1.8%) |  |
| Race | Caucasian | 1820 (98.3%) | 31 (1.7%) | 0.82 |
|  | African American | 106 (99.0%) | 1 (1.0%) |  |
|  | Asian or Pacific Islander | 24 (100.0%) | 0 (0.0%) |  |
| State | AL | 340 (98.8%) | 4 (1.2%) | 0.06 |
|  | FL | 885 (98.1%) | 17 (1.9%) |  |
|  | KY | 158 (96.3%) | 6 (3.7%) |  |
|  | Other | 574 (99.1%) | 5 (0.9%) |  |
| Education | ≤12 years | 652 (98.2%) | 12 (1.8%) | 0.85 |
|  | >12 years | 1183 (98.3%) | 20 (1.7%) |  |
| Hemisphere | Right | 934 (98.2%) | 17 (1.8%) | 0.86 |
|  | Left | 941 (98.5%) | 14 (1.5%) |  |
|  | Bilateral | 35 (97.2%) | 1 (2.8%) |  |
| Histology | GBM | 1245 (99.4%) | 8 (0.6%) | **<0.01** |
|  | Grade III | 323 (98.5%) | 5 (1.5%) |  |
|  | Grade II | 389 (95.3%) | 19 (4.7%) |  |

\*Unequal column totals reflect missing data

**Table 3** presents results from multivariable logistic regression considering factors associated with IDG. WHO grade II was associated with an over seven-fold increased prevalence of IDG (OR: 7.53; 95% CI: 2.87, 19.7) while controlling for gender and age. Neither age nor gender was independently associated with IDG after adjustment for tumor grade.

**Table 3.** Multivariate regression of clinical variates and asymptomatic presentation (N=1989)

| Category | Exposure | No. asymptomatic | OR* | 95% CI* | p-value |
| --- | --- | --- | --- | --- | --- |
| Tumor Grade | GBM | 8 | referent | - | - |
|  | Grade III | 5 | 2.41 | (0.75, 7.68) | 0.14 |
|  | Grade II | 19 | **7.53** | **(2.87,19.75)** | **<0.01** |
| Gender | female | 11 | referent | - | - |
|  | male | 21 | 1.47 | (0.70, 3.08) | 0.31 |
| Age at Dx | <50 | 20 | referent | - | - |
|  | >=50 | 12 | 0.96 | (0.42, 2.23) | 0.93 |

\*OR= odds ratio; CI=confidence interval. ORs/CIs adjusted for other terms included in the table.

Results of conditional Cox proportional hazards regression of IDG in relation to tumor-related death are shown in **Table 4**. HRs were estimated among 28 IDG patients and 63 individually matched symptomatic patients. (No suitable match could be identified in 4 of the 32 IDG patients.) As shown in Table 4, the hazard of dying was significantly reduced by ∼50% (HR: 0.49; 95% CI: 0.26, 0.95; p=0.035) in IDG adjusting through matching for age at diagnosis, date of diagnosis, and tumor subtype. Reduction in death rates was more pronounced when restricting to WHO grade II glioma (17 IDG and 36 symptomatic patients) (HR: 0.18; 95% CI: 0.03, 1.01; p=0.051) though results were based on limited data (1 death occurring among 17 IDG patients).

**Table 4.** Association of asymptomatic presentation with survival in matched analysis

| Parameter | HR | 95% CI | P-Value |
| --- | --- | --- | --- |
| IDG vs. symptomatic gliomas<sup>1</sup> | 0.49 | (0.26, 0.95) | 0.0349 |

Hazard ratio (HR) and 95% confidence interval (CI) for incidental glioma diagnosis (IDG) adjusted via matching for age at diagnosis (within 5 years), date of diagnosis (within 18 months), and tumor subtype (grade II, III or GBM). A total of 28 IDG and 63 individually-matched symptomatic patients were included in the analysis (no suitable match could be identified for the remaining 4 IDG patients).

In 14 of 28 matched sets, the IDG patient and controls were concordant for resection (N=**11 sets**) or biopsy-only (N=**3 sets**) whereas the remaining 14 sets were discordant for extent of surgery (in 12 of the 14 sets, the IDG patient underwent resection whereas the controls underwent biopsy-only). When restricting to the 14 matched sets concordant for surgery (comprised of 6 GBMs, 3 grade III gliomas, and 5 grade II gliomas), HRs were attenuated and no longer statistically significant (HR: 0.85; 95% CI: 0.42, 1.72; p=0.212). Among the 5 matched sets comprised of WHO grade II gliomas, 0/5 IDG and 5/11 symptomatic patients died from glioma during the observation period.

## DISCUSSION

The long-running clinic-based epidemiologic study, GliomaSE, offers a unique opportunity to describe the prognosis and natural history of subsets of patients with glioma. These subsets may offer insights as well as the generation of new hypotheses related to the course of these diseases. The study population is reflective of the population of patients with primary glioma diagnosed in the southeastern United States and genotyping of the study population has yielded results ^15,23^ in concordance with published glioma GWAS ^24^.

In the present study, we observed that as many as 5% of grade II gliomas are asymptomatic at presentation. Tumor grade is an overwhelming predictor of IDG, with a 7.5-fold excess of IDG in grade II tumors and nonsignificant 2.4-fold excess in grade III tumors when compared to glioblastoma. IDG was unrelated to patient age or gender in multivariate regression analyses after adjustment for tumor grade. The present study raises the possibility that IDG is associated with a longer survival interval following presentation among grade II tumors only although only tentative conclusions can be reached given the limited number of IDG patients available for analysis.

Previous studies of IDG included tumors that came to attention during workup for headache ^18,25- 27^. We elected to exclude such cases as there was no instance in our series of a tumor detected in persons with a long-standing headache disorder during a purely ‘routine’ imaging study. Rather, headaches leading to diagnosis were described by such persons as acutely worsening or of a ‘different character’ as compared to headaches usually suffered by the patient and this in turn prompted diagnostic work-up and detection of the tumor. We therefore classified such patients as ‘symptomatic’.

The study had some limitations. In spite of the large patient series, suitable matches in the survival analyses were not identified for several of the IDG patients (4 of 32). Furthermore, only 14 of the remaining 28 sets had controls matched also for tumor resection, a positive prognostic factor when compared to biopsy only ^11,12,28^. When restricting analysis to these 14 matched sets concordant for surgery type (all members of the matched set undergoing resection or biopsy), reduced mortality in IDG patients was greatly attenuated (HR=0.85) (not shown). We note that the latter result was based predominately on higher grade tumors (9 of 14 pairs had a grade III astrocytoma or GBM), and it is possible that earlier detection via incidental diagnosis is associated with survival benefit only in low-grade tumors: as noted, among the 5 matched sets comprised of WHO grade II gliomas, 0/5 IDG and 5/11 symptomatic patients died from glioma during the observation period. Of note, a recent study published by Ius *et al* examining incidentally discovered low-grade gliomas did show that incidentally detected low grade gliomas had a longer overall survival than symptomatic low grade gliomas, even when a complete surgical resection was obtained ^27^.

For most patients in this series, we lacked information on IDH mutation status which is a key prognostic feature of the low-grade tumors that comprised the majority of IDG patients in this study. (Most patients were diagnosed prior to the modern *IDH1* era). *IDH1* has been linked to improved patient survival^8,29,30^. In the present survival analysis, 39 of 91 patients had known *IDH1* status, and mutated *IDH1* was observed in a slightly higher proportion of IDG patients (52.9%, n=9 of 17) than non-IDG controls (40.9%, n=9 of 22) (p=0.43); among patients with WHO grade II tumors, 6/9 IDG and 4/10 symptomatic tumors were positive for *IDH1*.

Incidentalomas in GBM are rare and the potential impact on patient survival is poorly studied. Restricting to the 6 matched sets (5 GBM and 1 grade III astrocytoma) in which all members underwent resection within 3 months of tumor detection (mean: 27 days from presentation to surgery), IDG patients survived no longer (mean of 19.6 months; n=6) than the symptomatic patients (mean of 24.4 months; n=16); though based on limited data, these results suggest that incidental detection of the tumor does not extend survival in patients with high grade tumors.

The literature suggests that early identification of low-grade glioma is associated with improved outcomes as it is easier to operate on a smaller lesion ^10,31,32^. In addition, by detecting tumor prior to the onset of neurological signs and symptoms the burden of disease is the least on a patient and the opportunity for preserved neurological function remains the highest. The difference in survival outcome in this report supports efforts to identify risk factors associated with glioma. There is increasing support for a moderate role of genetic susceptibility through the identification of risk alleles ^24^. As further efforts define the biological significance of known risk variants and potentially expand this pool, genetic risk screening with targeted imaging may provide improved outcomes for patients, particularly the young adult with a low-grade glioma. This study provides some additional support for the developing evidence favoring early detection programs, “tailored screening,” and active management ^33^ which may include consideration of surgical resection, chemotherapy, radiotherapy, along with molecular and histopathologic characterization.

## Data Availability

The data analyzed for this manuscript are not available for dissemination.

